# An end-to-end platform for digital pathology using hyperspectral autofluorescence microscopy and deep learning based virtual histology

**DOI:** 10.1101/2023.04.10.23288259

**Authors:** Carson McNeil, Pok Fai Wong, Niranjan Sridhar, Yang Wang, Charles Santori, Cheng-Hsun Wu, Andrew Homyk, Michael Gutierrez, Ali Behrooz, Dina Tiniakos, Alastair Burt, Rish K. Pai, Kamilla Tekiela, Po-Hsuan Cameron Chen, Laurent Fischer, Eduardo Bruno Martins, Star Seyedkazemi, Daniel Freedman, Charles C. Kim, Peter Cimermancic

**Affiliations:** Verily; work done at Allergan; work done at Google; Newcastle University; Mayo Clinic; University of Athens

## Abstract

Conventional histopathology involves expensive and labor intensive processes that often consume tissue samples, rendering them unavailable for other analysis. We present a novel end-to-end workflow for pathology powered by hyperspectral microscopy and deep learning. First, we developed a custom hyperspectral microscope to non-destructively image the autofluorescence of unstained tissue sections. We then train a deep learning model to use the autofluorescence to generate virtual histological stains, which avoids the cost and variability of chemical staining procedures and conserves tissue samples. We showed that the virtual images reproduce the histological features present in the real stained images using a randomized nonalcoholic steatohepatitis (NASH) scoring comparison study where both real and virtual stains are scored by pathologists. The test showed moderate to good concordance between pathologists’ scoring on corresponding real and virtual stains. Finally, we developed deep learning-based models for automated NASH clinical research network (NASH CRN) score prediction. We showed that the end-to-end automated pathology platform is comparable to pathologists for NASH CRN scoring when evaluated against the expert pathologist consensus scores. This study provides proof of concept for this virtual staining strategy, which could improve cost, efficiency, and reliability in pathology, and enable novel approaches to spatial biology research.

## Introduction

Histological analysis typically involves the tissue biopsies, preparation (including fixation) of stained tissue sections (typically a few microns), followed by pathologist review. This process has been in use in some form for over 150 years and is considered the gold standard for tissue-based diagnostics. However, the process has the following limitations: first, the histological staining is laborious and expensive, especially for rare stains. The requirement of chemical reagents and degrading effects of age and environmental exposure on tissues impose additional costs and constraints. Second, staining often imposes irreversible changes on the tissue, precluding assessments on multiple biomarkers. This can become restrictive in cases with limited tissue access and prevents using the tissue for additional testing and research such as spatial biology analysis. Finally, staining, imaging and pathologist review processes may have high variability across different labs and facilities, which can impact clinical decision-making [1].

Recent advances in imaging technology and computer vision have spurred efforts to apply novel techniques to unstained tissue. These techniques include optical coherence tomography [2], which provides only structural information, nonlinear microscopy accompanied by nuclear staining [3], fluorescence lifetime imaging [4], and vibrational spectroscopy techniques including infrared imaging [5], stimulated Raman scattering [6], coherent anti-Stokes Raman scattering [7] and multimodal combinations [8] that maximize molecular information at the expense of spatial resolution or imaging speed. In comparison, multispectral and hyperspectral autofluorescence (AF) imaging [9, 10] allow distinction of several endogenous fluorophores at high spatial resolution using relatively simple optical setups with scan speeds suitable for whole-slide imaging. They also enable implementations with parallelized detection across emission wavelengths to minimize photobleaching of the sample.

However, these novel imaging methods must be evaluated against the current gold standard of manual assessments by pathologists using brightfield (BF) microscopes on chemically stained tissue. Furthermore, it would take substantial additional training for pathologists to be familiar with these alternative imaging outputs. On the other hand, new deep learning techniques have been shown to assist pathologists with improved diagnostic efficiency and accuracy on BF images [11]. While such methods are easier to incorporate in the conventional pathology workflow [16], they only accelerate the final diagnostic step of the workflow and do not address the drawbacks in the staining process itself. Thus, there exists an opportunity to bridge the gap between the emerging technologies in imaging and machine learning (ML) and the deep clinical expertise of pathologists using stained brightfield images for diagnosing diseases and assessing their severity.

Here, we present a combined imaging and staining digital pathology platform that digitizes unstained tissue sections and then uses deep learning to perform virtual staining (Figure 1). The tissue digitization step in our workflow is based on AF imaging of unstained tissue at hundreds of excitation-emission wavelength pairs using our hyperspectral microscope. The virtual staining step utilizes recent advances in computer vision and deep learning. Importantly, the platform is non-destructive, with the tissue samples remaining unstained and thus available for other analyses.

**Figure 1.**
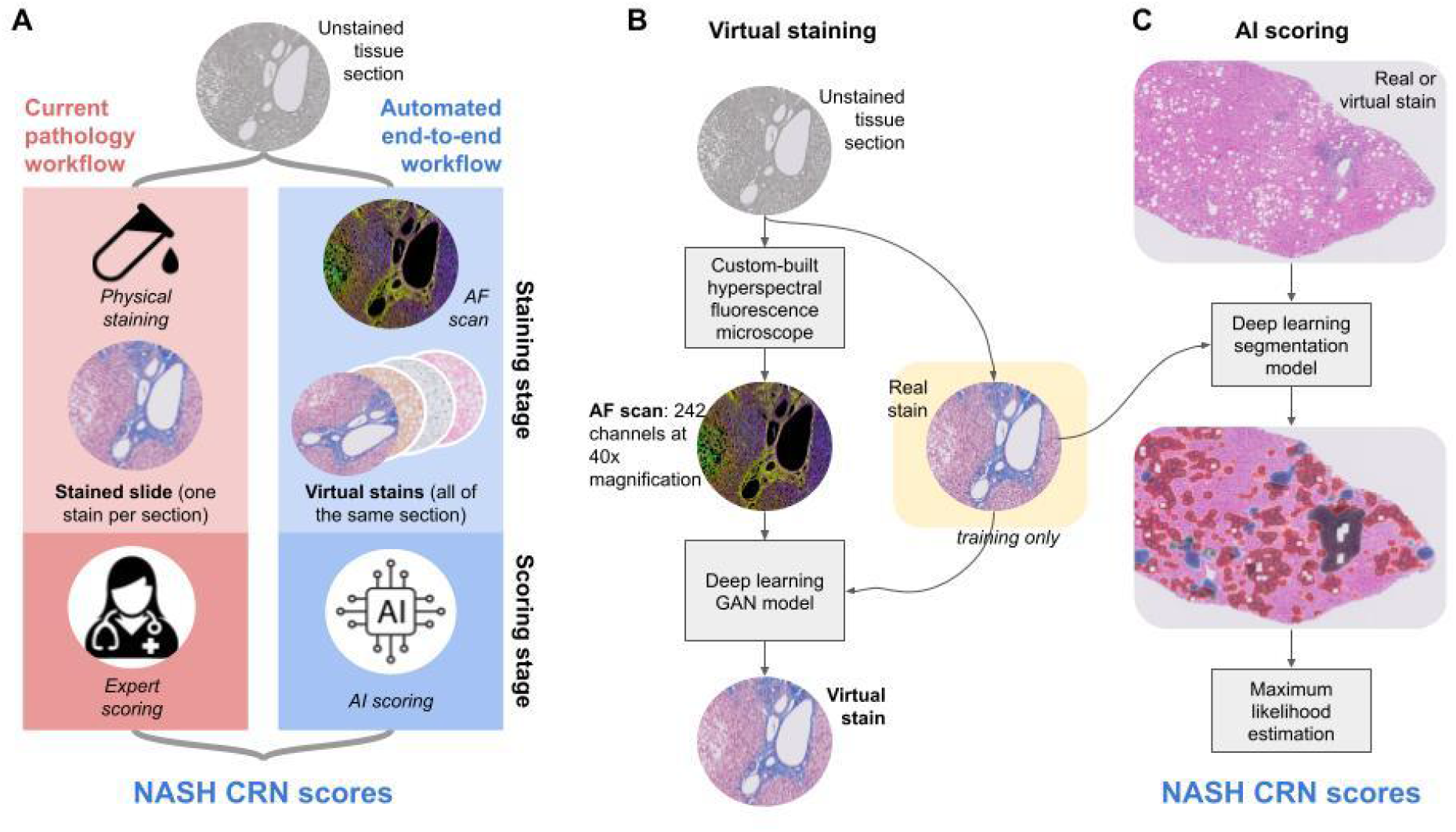
a) We present an automated workflow with virtual staining and AI scoring that mimics the steps of the current pathology workflow. b) The Virtual Staining pipeline uses a custom hyperspectral microscope to image unstained tissue samples. Our deep learning virtual stainer model takes AF images as input and returns stains images that are trained to look like real chemically stained tissue samples. c) Our AI scoring models use BF images of chemically or virtually stained tissues to estimate the presence of NASH features and NASH CRN scores.

To evaluate the platform, we applied it to tissue sections from a multi-arm clinical study on nonalcoholic steatohepatitis (NASH) [12], and evaluated it using both a hepatopathologist scoring comparison study and artificial intelligence (AI) scoring algorithms. NASH is a progressive form of non-alcoholic fatty liver disease (NAFLD) that affects over 15 million people in the US annually [13]. The histologic features of NASH include macrovesicular or mixed steatosis, hepatocellular ballooning and Mallory-Denk bodies (MDBs), scattered (mainly lobular) inflammation, and fibrosis [12]. As no single feature of NASH is diagnostic by itself, the NASH Clinical Research Network system (NASH CRN) was devised for assessing the severity of NASH [14]. The NASH CRN system uses specific features that are identifiable on liver tissue stained with hematoxylin and eosin (H&E) and a collagen stain, usually Masson’s trichrome (MT). H&E differentially pigments the tissue, coloring the nuclei, cytoplasm, other cell structures and the extracellular matrix with different shades of pink and purple. MT stains the extracellular matrix and highlights the amount and distribution of fibrosis. In a typical NAFLD diagnostic workflow, hepatopathologists evaluate a panel of stained slides (including H&E and MT) from a liver biopsy and, after making the diagnosis, quantify the histological features using the NASH CRN system. The NASH application enabled a comprehensive evaluation of our platform by involving two different stains, multiple histologic features, and a specific scoring system.

## Results

### Hyperspectral Microscopy

We built a hyperspectral microscope to probe the near-ultraviolet through the near-infrared portion of the excitation-emission space of the unstained tissue. To reduce photobleaching and minimize scan time, the microscope used a parallelized detection scheme to collect multiple wavelengths at the same time and make efficient use of the collected light. More importantly, it provided finer spectral resolution than required to resolve known AF features such as collagen, elastin, NADH, flavins, lipopigments, and porphyrins [15]. Figure 2 illustrates the spectral information contained in an example AF image. The images in the bottom row (Figure 2, G-J) were computed as linear projections from a single hyperspectral image. The projection vectors (Figure 2B) were determined by Canonical Correlation Analysis, to maximize the correlation of the projected images across the entire slide with real immunofluorescence signals. We observed different tissue features with different spectral responses, and the projected images showed morphological similarities with the real-stained images shown in Figure 2 (C-F). Averaging over a limited spectral band such as those captured by conventional AF imaging microscopes did not allow for easy differentiation between different tissue features, while spectral projections of the hyperspectral image could differentiate features such as extracellular matrix, nuclei, and macrophages.

**Figure 2.**
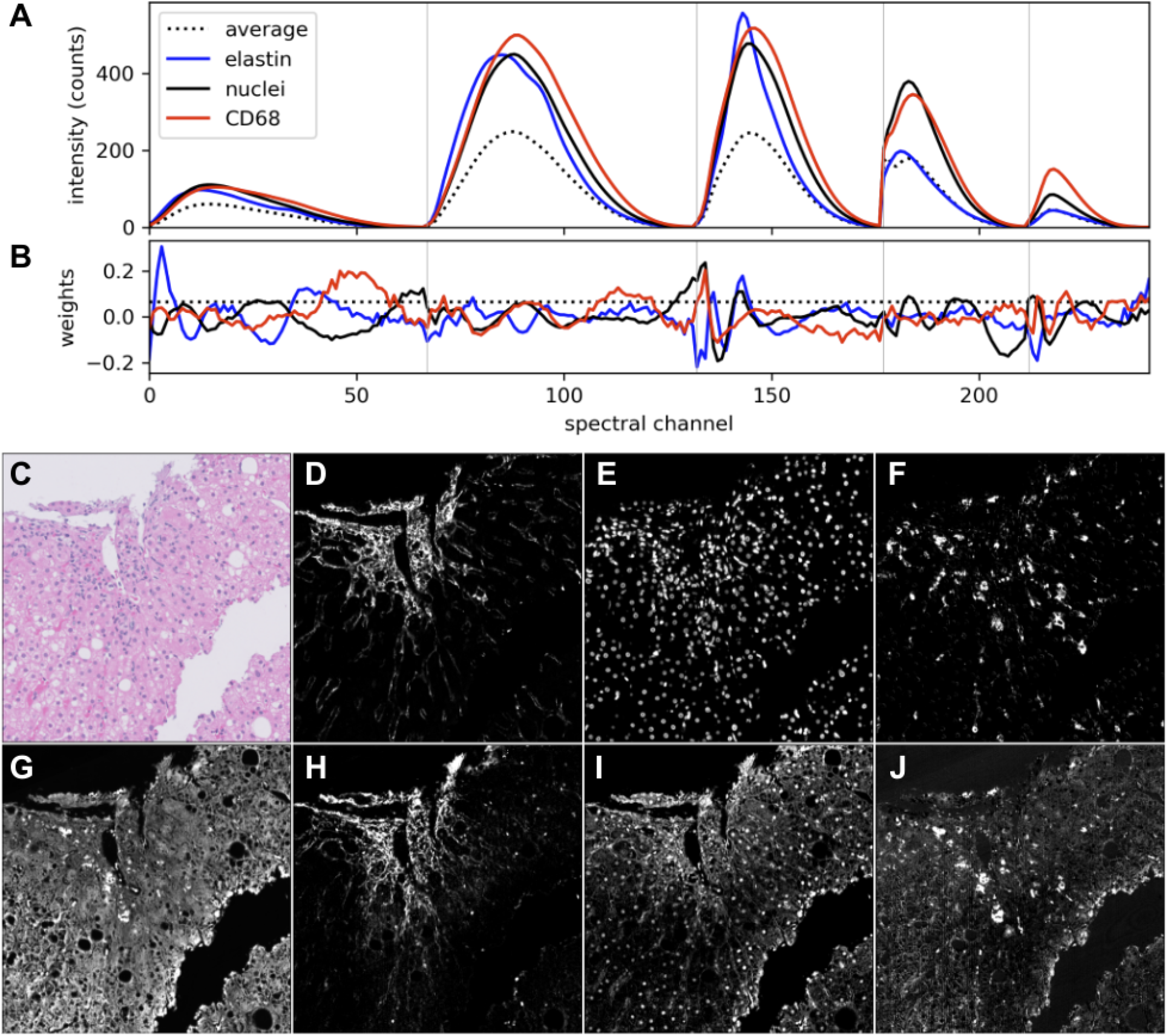
(a) Masked, weighted-average AF spectra computed for elastin, nuclei and CD68-containing regions on a single FFPE liver biopsy section. The vertical lines separate channels corresponding to the five different excitation lasers. (b) Weights used to generate the spectral projection images below. (c) Real H&E stained image. (d-f) Real immunofluorescence images of (d) elastin + α-SMA, (e) nuclei, and (f) CD68. (g-j) Spectral projections computed from the hyperspectral AF image: (g) a uniform projection (sum across all spectral channels) showing general tissue features including cytoplasm, (h) a projection enhancing extracellular matrix components, (i) a projection enhancing nuclei, and (j) a projection enhancing macrophages and lipofuscin.

### Virtual Stainer

We trained a virtual stainer model (Figure 3) that can produce high quality virtually stained BF images using AF images (Figure 4). The model is based on pix2pix style image translation architecture [20].

**Figure 3.**
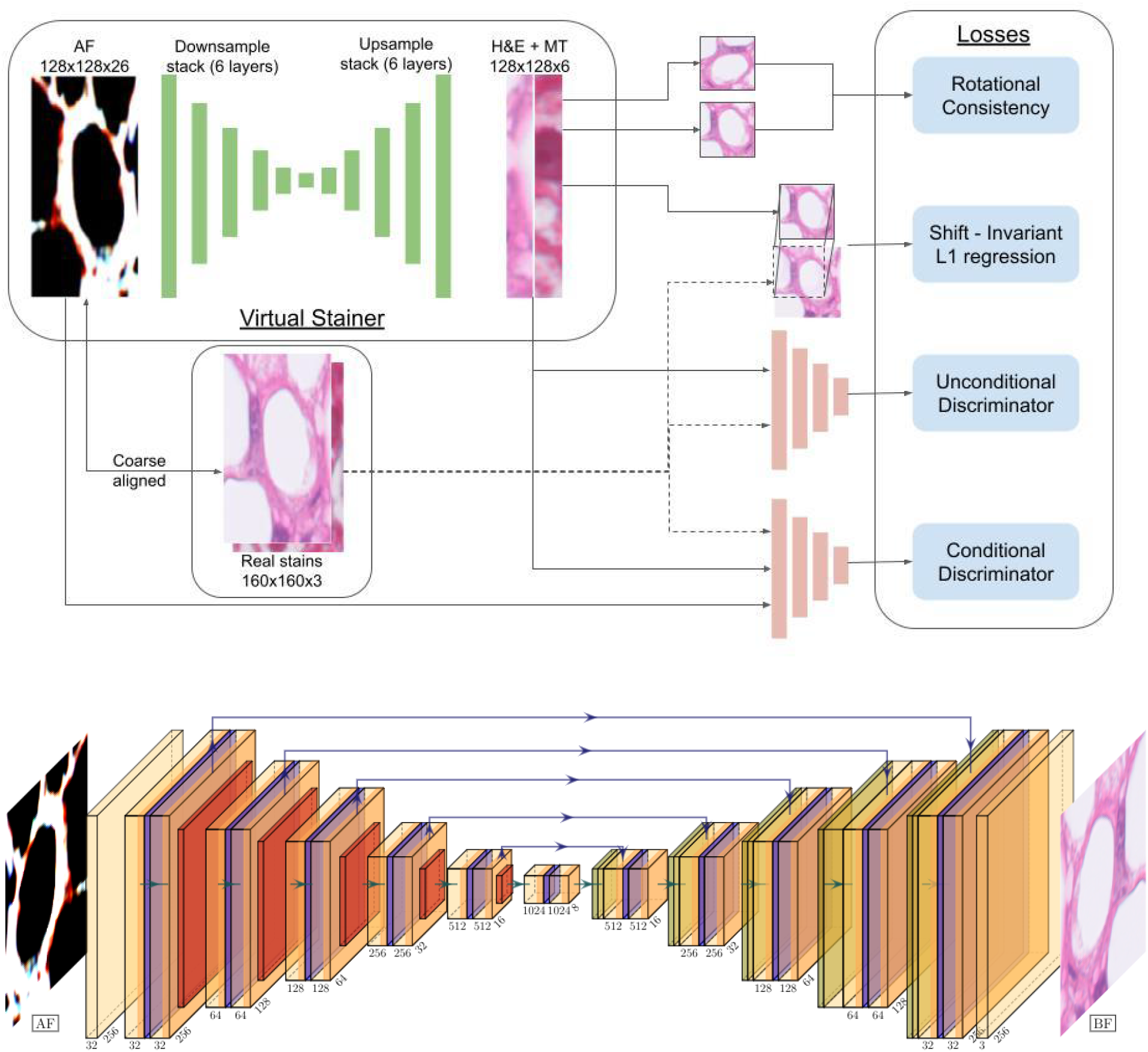
a) Virtual stainer training pipeline. The virtual stainer is a neural network with a Unet architecture. It takes patches of AF images of size 128x128 and returns 2 images - 1 H&E and 1 Trichrome BF image. We use chemically stained BF images aligned to the AF images to train the virtual stainer using four different training losses. b) The neural network architecture of the generator of virtual stainer.

**Figure 4.**
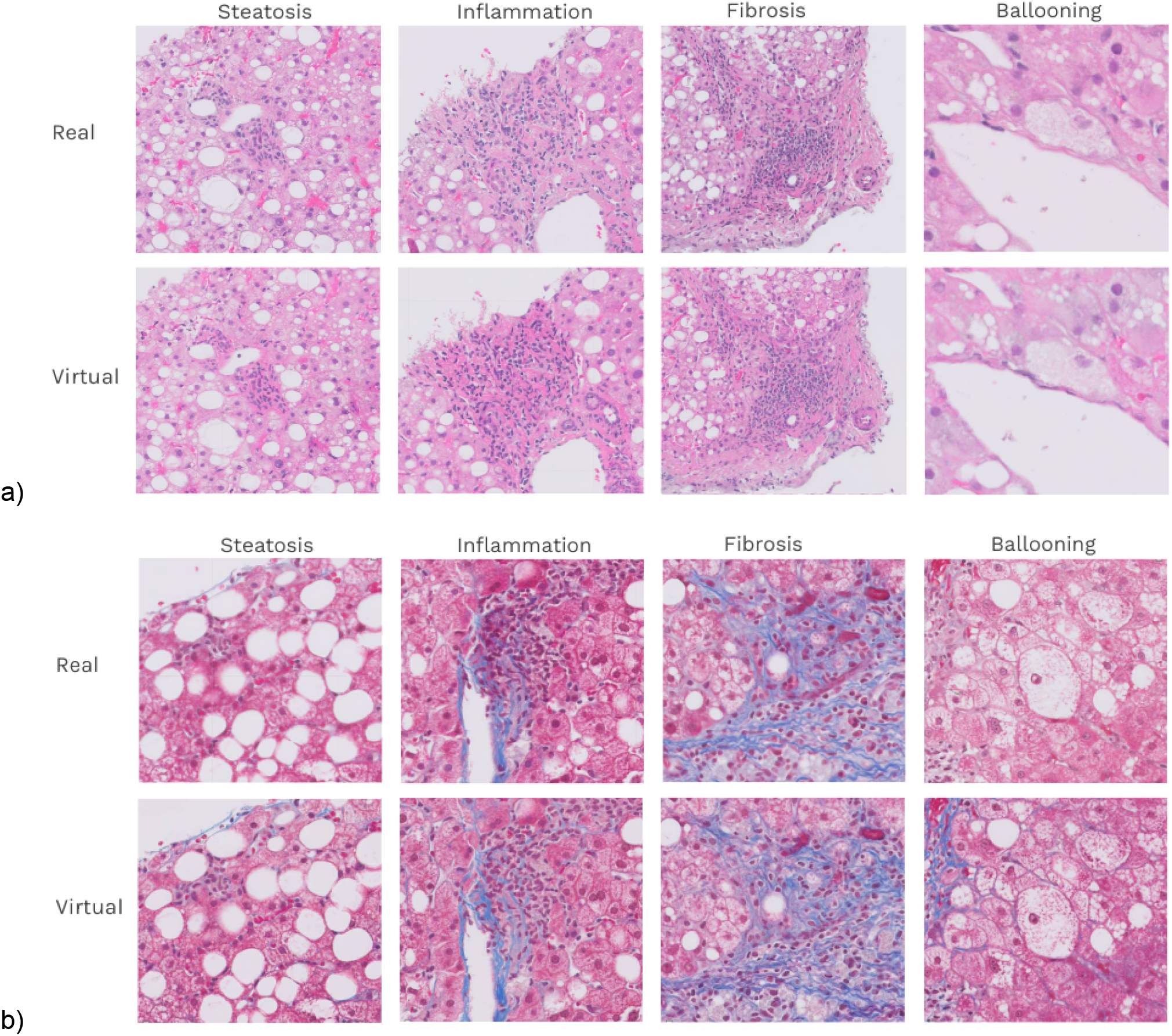
Comparing real and virtual stains at 40X magnification shows that virtual stains accurately capture both the spectral and morphological attributes of real stains. Here we show examples of chemically stained patches which show the 4 characteristic NASH features and their corresponding virtual stain a) BF real and virtual images stained with H&E b) BF real and virtual image stained with MT.

We evaluated the virtual stains using a number of independent criteria which can be divided into 3 parts: (1) image quality, (2) automated NASH feature segmentation, (3) automated NASH CRN scoring, and (4) human-expert comparison study using NASH CRN scoring.

#### 1. Image quality metrics

To evaluate the quality of the images produced by Virtual Stainer algorithmically, we used the brightfield microscope image as the reference (ImgR) and the virtual stainer image as the prediction (ImgP) and measured various metrics to capture the difference between the two.

As seen in Table 1, our virtual stainer ranks between fair to good on all image quality metrics. In particular we note that our SSIM (which captures the structural information that humans look for in an image) is the highest reported number in virtual staining literature [21].

**Table 1:** Image reconstruction measures (absolute error, mean squared error, peak signal-to-noise ratio and structural similarity index measure) were calculated using the real stains as reference. Each metric is aggregated over the whole slide images ranging from 5000 to 50000 pixels.The values in the table are the slide level metrics averaged over all the slides and the values in the parentheses indicate the standard deviation of the metrics over all the slides.

| Metric | H&E | Masson's Trichrome |
| --- | --- | --- |
| Relative mean absolute error | 5.7 % (0.7) | 6.6 % (1.4) |
| Relative root mean squared error | 8.2 % (1.6) | 8.7 % (1.8) |
| PSNR | 35.7 (0.6) | 35.0 (0.9) |
| SSIM | 0.98 (0.04) | 0.98 (0.01) |
| Num whole slides | 73 | 40 |

#### 2. Automated NASH feature segmentation

The NASH segmentation models (Stage 1 of our NASH scoring suite) return one summary attribute value per NASH feature per slide quantifying the presence of NASH histologic features in the whole slide. The models were patch level image recognition networks based on InceptionV3 architecture [19].

We can evaluate the quality of the segmentation model predictions on the independent TEST set (94 slides for H&E and 59 slides for MT) by measuring the correlation of slide-level attributes with the pathologist-assigned NASH CRN scores (Figure 5). The segmentation models reach the spearman correlation values of 0.76, 0.50, 0.53, 0.64 for steatosis, lobular inflammation, ballooning and fibrosis, respectively, comparable with those in previous studies [22]. All 4 correlations had p-values less than 1e-5.

**Figure 5.**
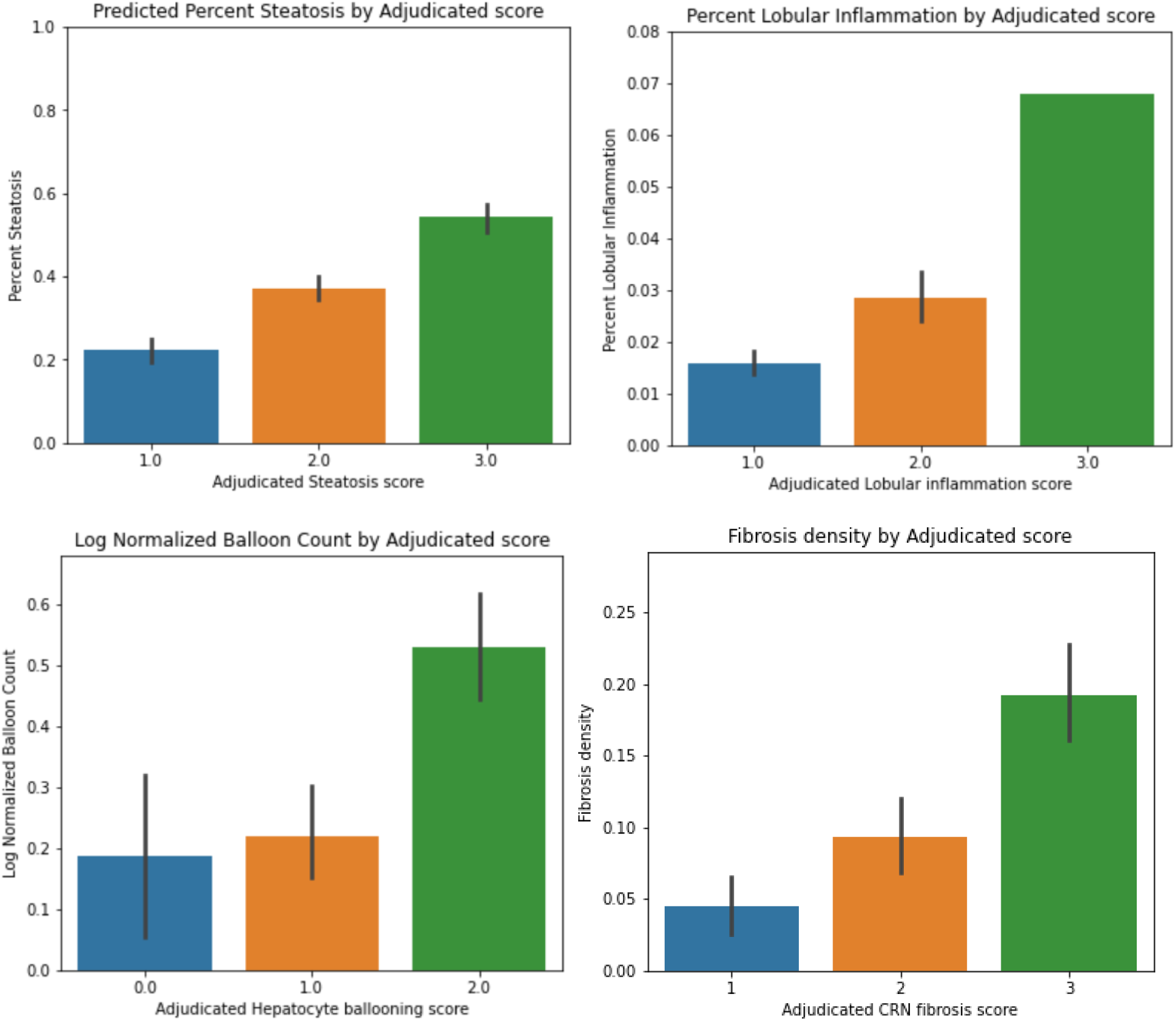
Correlation of slide level NASH feature attributes predicted by segmentation models on real stains with NASH CRN scores adjudicated by hepatopathologists. In this figure, we pick 1 attribute for each of the 4 NASH features - a) percent steatosis b) percent lobular inflammation c) log normalized balloon count and d) fibrosis density.

We used the segmentation models to evaluate the quality of virtual stains by comparing the predicted summary attributes of real stains and their corresponding virtual stains. If the virtual stains have successfully captured the relevant NASH features present in the real stains, then we can expect the segmentation models that were exclusively trained on real stains to have similar predictions on the real and virtual stains of the same slide.

The steatosis segmentation model predicted the fraction of the whole slide with steatosis (Figure 6A). Therefore, we plotted the fraction of steatosis predicted for the same tissue sample, with real slide on the x-axis and virtual slide on the y-axis. Similarly, for lobular inflammation, we plotted the predicted fraction of the whole slide with lobular inflammation (Figure 6B). For ballooning, we plotted the log of the number of ballooning cells in the whole slide normalized by the size of the slide (Figure 6C). Finally, the fibrosis segmentation model predicted patch level presence of fibrosis, hence we plotted the fraction of patches predicted to contain fibrosis in real and virtual stains (Figure 6D). We observed Pearson correlation values of 0.95, 0.81, 0.89, 0.53 for steatosis, lobular inflammation, ballooning and fibrosis, respectively with all p-values being less than 1e-5.

**Figure 6.**
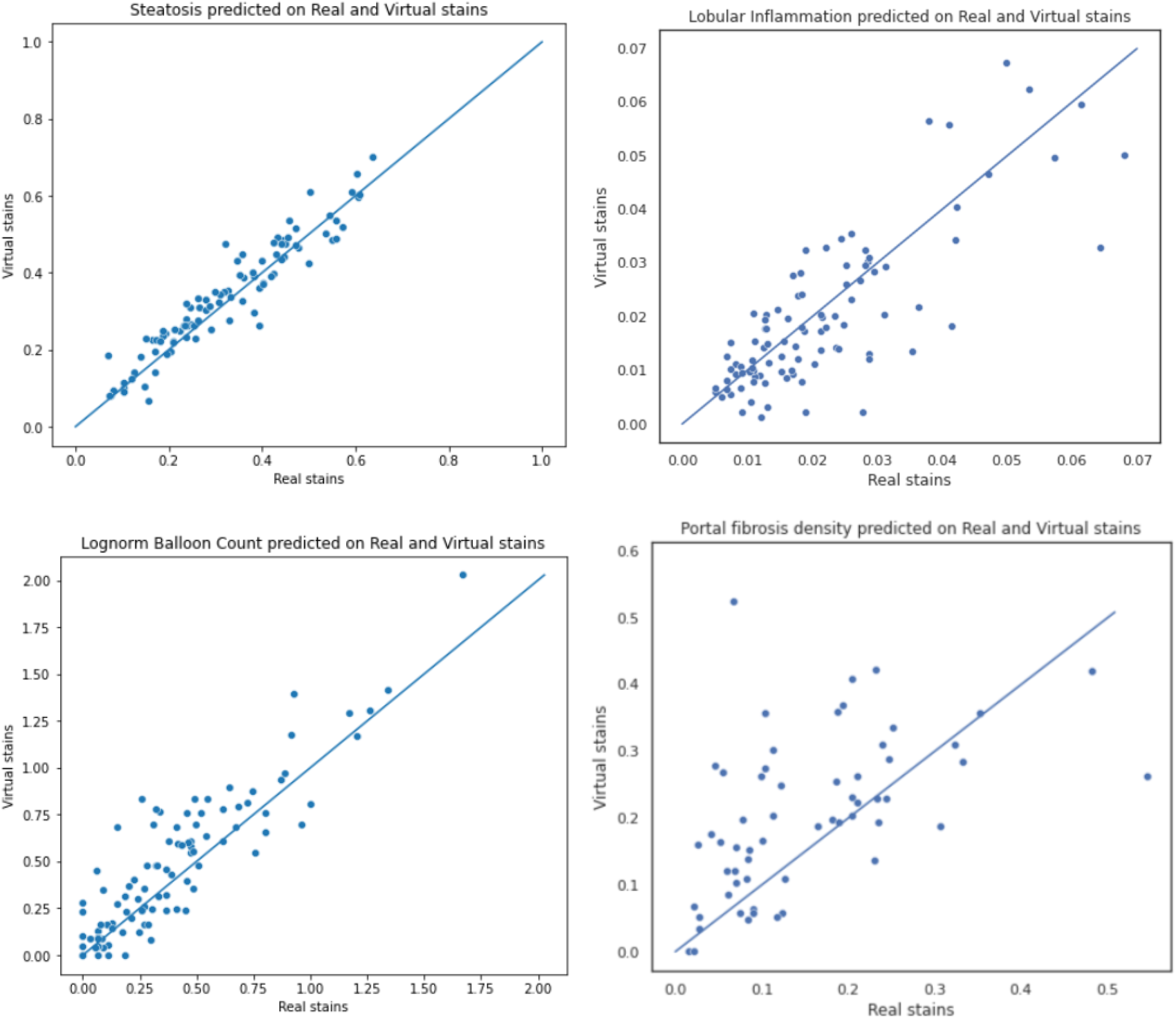
Correlation of slide level NASH feature attributes predicted by segmentation models on real stains vs virtual stains. In this figure, we pick 1 attribute for each of the 4 NASH features - a) percent steatosis b) percent lobular inflammation c) log normalized hepatocyte balloon count and d) fibrosis density.

#### 3. Automated NASH CRN Scoring

The Stage 2 scoring models used the output of the Stage 1 segmentation models to predict NASH CRN clinical scores for each NASH feature.

We evaluated the models’ predicted scores against the harmonized NASH CRN scores applied by the adjudicators on our independent test set, using linearly-weighted Cohen’s kappa as the metric of evaluation. The test set had 94 H&E slides used for scoring steatosis, ballooning and lobular inflammation and 58 MT slides used for scoring fibrosis. Finally, we compared Cohen’s kappa measures achieved by our scoring suite on real stains, our scoring suite on virtual stains and independent hepatopathologists on real stains. We report the kappas for all three comparisons, along with their bootstrapped 90% confidence intervals (CI) in Table 2.

**Table 2.**
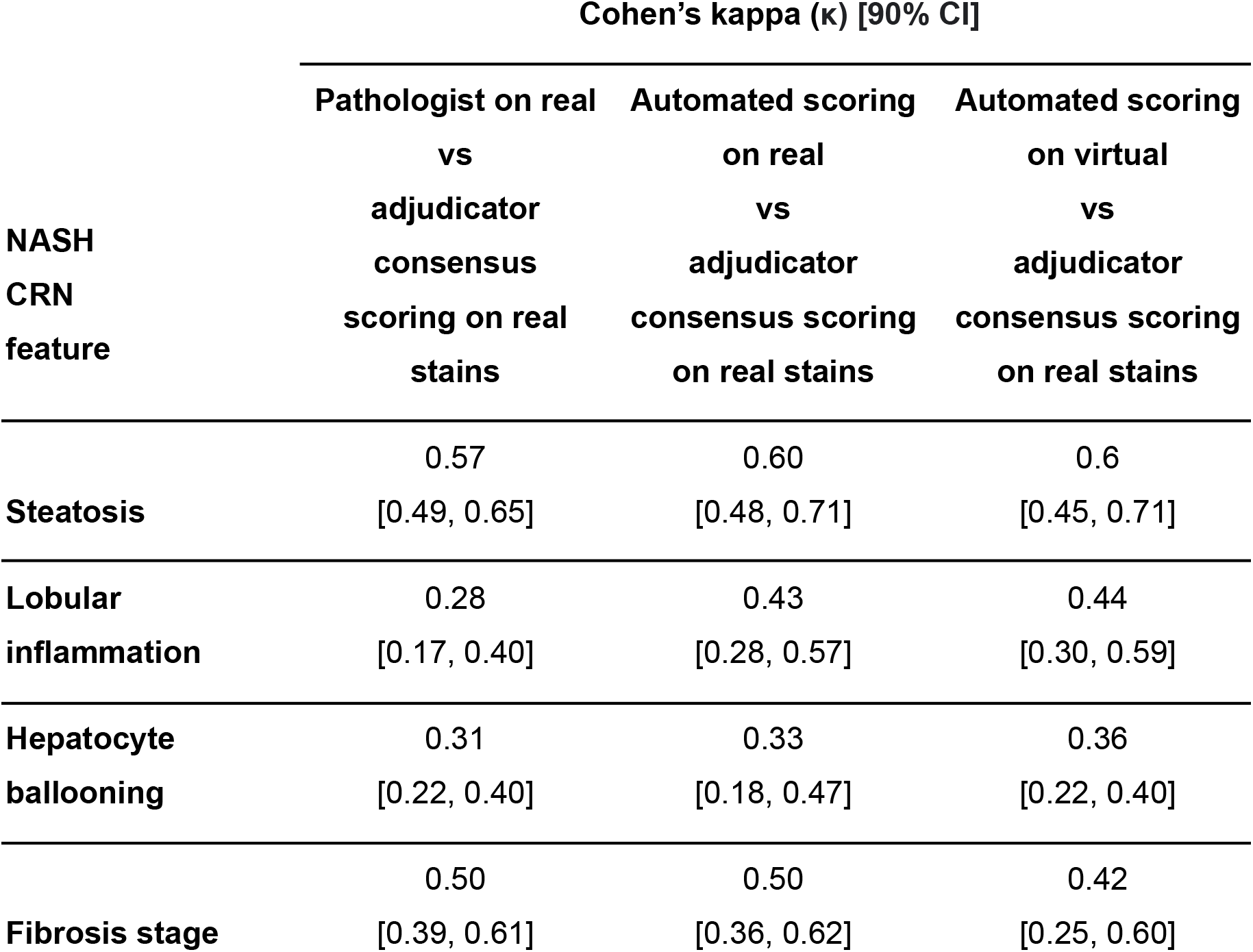
We compare the scoring of a) hepatopathologists on real stains, b) automated scoring on real stains, and c) automated scoring in virtual stains, against the adjudicator pathologist consensus scores. We show the linearly-weighted Cohen’s kappa values of each comparison for each of the 4 NASH features along with the 90% confidence intervals (CI) estimated using bootstrap.

#### 4. Human Expert Comparison Study using NASH CRN Scoring

Finally, we performed a comparison study to directly measure the difference in NASH CRN scoring done by pathologists when they score chemically stained BF images versus virtually stained images.

Three experienced hepatopathologists, each with 15-30 years of experience, scored NASH features for both real and virtual H&E and MT stains. The pathologists arrived at a consensus scoring for each slide. For H&E slides, they scored 3 NASH features - steatosis, lobular inflammation and ballooning - and for MT slides, they scored 1 NASH feature - fibrosis.

The pathologists reviewed a total of 180 slides, 45 pairs of H&E stains and 45 pairs of MT slides shown in random order with a washout period between real and virtual staining modalities of at least 1 week. Each pair is made up of a real, chemically stained BF image and virtually stained image predicted from the AF image of the same tissue sample. Thus for each tissue sample, we obtained 2 sets of NASH scores - one scored on real stains and one scored on virtual stains.

We then ascertained the concordance (linearly weighted κ) between the real and virtual NASH score. As shown in Table 3, we observed moderate to high agreement between real and virtual NASH scores. Accuracy and kappa agreement was 0.91 (0.82, 0.98) and 0.86 (0.73, 0.96) for steatosis, 0.73 (0.62, 0.82) and 0.33 (0.06, 0.57) for lobular inflammation, 0.84 (0.75, 0.93) and 0.76 (0.61, 0.89) for ballooning and 0.65 (0.52, 0.77) and 0.55 (0.35, 0.70) for fibrosis.

**Table 3.** The accuracy and linearly weighted Cohen’s kappa of NASH CRN scoring performed by adjudicator pathologists on real stains versus virtual stains.

| NASH CRN feature | Accuracy [90% CI] | Cohen's kappa ($\kappa$) [90% CI] |
| --- | --- | --- |
| Steatosis | 0.91 [0.84, 0.97] | 0.86 [0.73, 0.96] |
| Lobular inflammation | 0.73 [0.62, 0.84] | 0.33 [0.06, 0.57] |
| Hepatocyte ballooning | 0.84 [0.75, 0.93] | 0.76 [0.61, 0.89] |
| Fibrosis stage | 0.64 [0.52, 0.75] | 0.55 [0.35, 0.70] |

We plot the distributions of kappa for each of the comparisons - pathologists on real stains, automated scoring on real stains and automated scoring on virtual stains, all measured against the adjudicator scoring (Figure 7).

**Figure 7.**
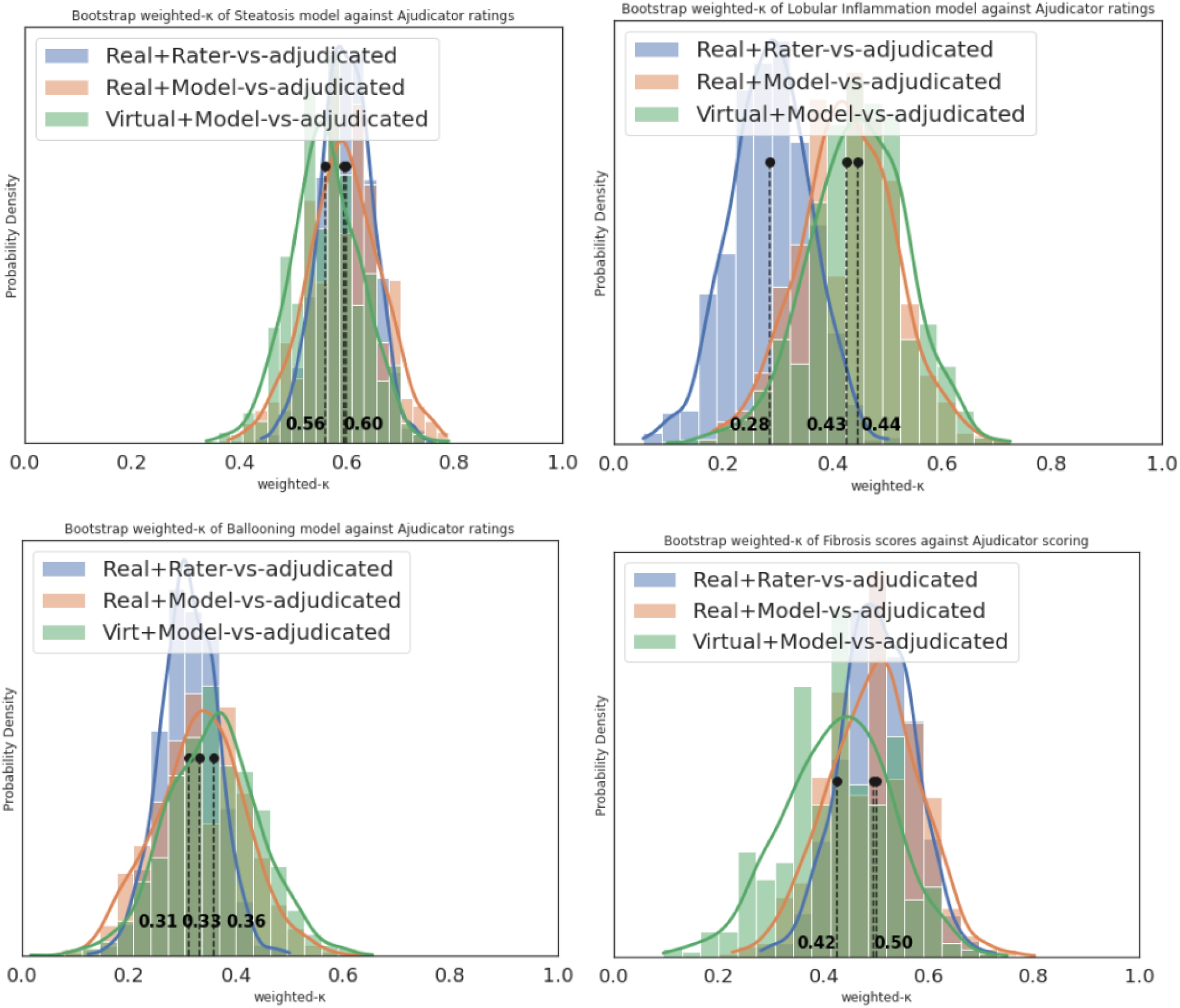
We plot the distributions of 1000 bootstrapped kappa estimates comparing estimated scores against the adjudicator consensus scores. The blue distributions are the weighted kappa of scoring performed by a randomly sampled hepatopathologist against the adjudicators reference. The red distributions are the kappa of our NASH scoring models applied on brightfield images of real stains. The green distributions are the weighted kappa of scoring by our end-to-end automated platform using virtual stains and automated NASH scoring models. The four plots show the four NASH features: a) steatosis, b) lobular inflammation, c) ballooning, d) fibrosis.

## Data Availability

The data for the patient samples cannot be shared under the restrictions placed by the institutional review board. The data storage and retrieval infrastructure was built and managed by Google LLC.

## Contributions

M.G. and C.S. led the development and calibration of the hyperspectral microscope and data collection of AF images. P.C., C.C.K., A.H., L.F., E.B.M., and S.S. led the study design. P.F.W. led the histopathology and BF image data collection, and designed protocols for pathologist annotations and scoring. P.F.W., P.C.,D.T., A.B., and R.K.P. led the NASH scoring data collection. K.T. and P.C.C. built and maintained the data infrastructure for storing, processing and retrieval of AF images and real and virtual BF images.

C.M., Y.W., P.C.C., N.S. and D.F. designed and wrote code for the data infrastructure, network architecture and training and testing pipelines of virtual stainer. C.M., Y.W., N.S. wrote code for the data infrastructure, network architecture and training and testing pipelines of NASH scoring models. C.M., Y.W., A.B. and N.S. performed analysis of NASH scoring models. A.H. and C.H.W wrote earlier versions of virtual staining model development pipelines.

N.S. wrote the manuscript with contributions from all authors. P.C. led the design and concept of the project and supervised all aspects of the projects. A.H., C.S., K.T., and C.H.W. worked on and wrote code for data acquisition, data ingestion, and earlier versions of model development and testing. P.F.W. advised and reviewed model iterations.

## Acknowledgments

We thank Allergan/AbbVie for their collaboration and sharing the CENTAUR clinical trial data [17,18]. We thank members of the translational pathology and hardware teams at Verily who supported tissue processing, quality checking, staining, and scanning: James Higbie, Hardik Patel, Julia Sigman, Robert Findlater, Vanessa Velez, Tzu-Chien Wang. We thank Dr. Sudha Rao and Dr. Debra Hanks for their pathology expertise, advice and contributions to various aspects of the work. We thank Susan Kram, Nina Lottsfeldt, and Janelle Chang Clark for program management support. We thank various members of Verily LIMS and lab engineering teams, including: Tyler Berry, Woody Ahern, Ian King, and Byron Boagert. We thank Dr. Thomas Snyder for early leadership and support.

